# Impact of Surgeon cadre, theatre location, and presence of intern healthcare professionals on decision-to-delivery interval in women undergoing emergency caesarean section in Northern Uganda: a historical cohort study

**DOI:** 10.1101/2024.12.16.24319122

**Authors:** Henry Ochola, Ronnie Omoro, Paul Buga, Emintone Ayella Odong, Oscar Ocaya, Rogers Kajabwangu, Emmanuel Ochola, Nelly Atim, Harriet Akello, Judith Praiselyn Acayo, Doris Ekwem, Jovia Namuddu, Derrick Mukurasi, Enock Lukyamuzi, Hudson Onen, Maurine Lenia, Martha Gimono, Emily Webb, Oona Campbell, Ronald Komata, Jerom Okot, Sande Ojara

**Author notes:** Corresponding author (HeO).

## Abstract

**Introduction:** Emergency caesarean sections (CS) are crucial for preventing life-threatening complications. The Decision-to-Delivery Interval (DDI), the time between decision and actual delivery, impacts maternal and neonatal outcomes. While the World Health Organization recommends a DDI of < 30 minutes, achieving this in low-resource settings remains challenging. This study examines the impact of surgeon’s cadre, operating theatre location, and presence of intern healthcare professionals on DDI and whether these associations vary by CS indication.

**Methods:** This historical cohort study was conducted at St. Mary’s Hospital Lacor, a tertiary hospital in Northern Uganda, involving 760 women who underwent emergency CS between 6^th^ September 2022 and 1^st^ June 2024. We assessed the association of prolonged DDI (≥60 minutes) with surgeon cadre, operating theatre location, and intern presence using logistic regression, adjusting for key confounders and investigated interaction with the indication for emergency CS.

**Results:** The median DDI was 51 minutes (IQR: 36-67), with 36.0% of cases classified as prolonged (≥ 60 minutes). Emergency CS performed by junior doctors had twice the odds of prolonged DDI compared to senior doctors (adjusted OR: 2.07; 95% CI: 1.38-3.10). Theatre location showed no effect on DDI (OR: 0.89; 95% CI: 0.61-1.28). The presence of interns was weakly associated, with slightly lower odds of prolonged DDI when interns were absent (OR: 0.71; 95% CI: 0.51-1.02). No significant variations were found based on the indication for emergency CS.

**Conclusion:** Surgeon’s cadre is a key factor in reducing prolonged DDI, highlighting the importance of training and supervision for junior doctors. While theatre location did not significantly impact DDI, improving theatre readiness and coordination remains essential. The weak association with intern presence suggests further investigation into their role in emergency CS. These findings highlight the importance of addressing system-level delays to improve timely emergency obstetric care in resource-limited settings.

## Introduction

Emergency caesarean section (CS) is a surgical procedure performed to prevent immediate life-threatening or life-limiting complications to the foetus and/or the mother (1). As a critical component of emergency obstetric care (EmOC), CS is offered when vaginal delivery is either challenging or absolutely impossible (2). The indications for CS are categorized using the four-grade classification system developed by Lucas et al., based on the urgency of the procedure (3). The Decision-to-Delivery Interval (DDI) refers to the time duration between the decision to perform an emergency CS and the actual delivery of the baby (4). Timely completion of emergency CS is crucial for optimal maternal and neonatal outcomes (5). While the World Health Organization (WHO) recommends a DDI of less than 30 minutes for emergency CS (6), this target is often challenging to achieve in low-resource settings due to various system challenges (7–9).

In developed countries, DDIs of 20 minutes or less are routinely achieved, largely due to better organizational structures (10,11). However, in resource-limited countries, factors such as delayed patient preparation (12), prolonged operating room preparation (13), and unavailability of operating rooms (14) contribute to extended DDIs. Interdisciplinary partnership and effective communication among team members have been shown to reduce DDIs (15). While some studies have shown no effect of prolonged DDI on foetal and/or maternal outcomes (16,17), others, particularly in Sub-Saharan Africa, have demonstrated associations between delayed emergency CS and increased maternal complications (18) and neonatal asphyxia (8,19). A study at Mulago National Referral Hospital in Uganda found an average DDI of 5.5 hours, and that increased DDI was associated with poor neonatal outcomes (20).

Because of insurgency and poverty, health facilities in Northern Uganda grapple with major challenges which are likely to lead to prolonged DDI. However, the DDI has not been quantified in these facilities and neither have the health-facility-related factors that influence it been studied. We aimed to study the effect of surgeon cadre, proximity of operating theatre to the ward, and presence of intern healthcare professionals on DDI in women undergoing emergency CS at a tertiary hospital in Northern Uganda and whether these associations vary by the CS indications.

## Materials and methods

### Study design and setting

We conducted a historical cohort study at St. Mary’s Hospital Lacor, a 482-bed private non-profit Catholic referral hospital located in Gulu City, Northern Uganda. The hospital primarily serves the Acholi, Lango, West Nile, and Karamoja subregions and South Sudan. The hospital’s Maternity unit, part of the Department of Obstetrics and Gynaecology, manages over 6,000 deliveries annually, including over 1800 CS, of which nearly 600 are emergencies (21).

The Maternity unit is staffed by three obstetricians, four medical officers, 28 midwives, and additional support staff. Emergency CS are performed by junior doctors (interns) or senior doctors (medical officers or obstetricians). Anaesthesia is provided by a team of 14 anaesthetists of varying qualifications, offering both spinal and general anaesthesia in both operating theatres. Despite the semi-consistent use of intrapartum monitoring tools, maternal and foetal monitoring in labour is done using the WHO labour care guides and cardiotocography by the midwives and doctors. Being a paid service, the payment for the emergency CS are made by the woman or her relatives after the emergency CS is performed.

### Participants

We retrospectively extracted data for women at ≥ 24 weeks of gestation admitted at the maternity unit of St. Mary’s Hospital Lacor who were prescribed an emergency CS by a doctor. The indications for the emergency CS included foetal distress, cord prolapse, placenta previa, abruption placenta, eclampsia, obstructed labour, failed vacuum extraction, or previous CS scars in active labour. We excluded participants with multiple pregnancies and gestational age <24 weeks.

### Data collection

We collected data for this study between 6^th^ September 2022 and 1^st^ June 2024, utilising records from a concurrent quality improvement project focused on enhancing documentation, tracking, and reporting of emergency CS. Data were extracted using a standardised collection tool to ensure consistency across all participants, regardless of exposure status.

### Variables

The primary outcome variable was the DDI, measured in minutes, defined as the time from the decision to perform an emergency CS to the time of delivery of the baby. DDI was categorised as prolonged (≥60 minutes) or not prolonged (<60 minutes) according to the local results-based financing guidelines(22).

The exposure variables were:

**i. Surgeon cadre performing the emergency CS -** Juniors or senior doctors (medical officers or obstetricians). We defined juniors as intern doctors who upon completion of their undergraduate study in medicine are deployed to tertiary hospitals by the Uganda Ministry of Health for hands-on training for one year before being fully licensed to practice; medical officers as fully licensed general medical doctors; and obstetricians as fully licensed medical doctors with specialised training in women’s reproductive health.
**ii. Operating theatre -** Emergency CS are performed in either the labour-suite theatre (operating 8 hours on weekdays) or a main theatre (105 meters -nearly 3 minutes’ walk from the labour-suite), functioning continuously with 1-2 operation tables dedicated to emergency CS). Additionally, the main theatre is preferred for potentially complicated CS due to its proximity to the intensive care unit.
**iii. Presence of intern health professionals –** Intern doctors, midwives, nurses, and pharmacists, deployed annually by the Ministry of Health in Uganda, add to the workforce at St. Mary’s Hospital Lacor. Given the inconsistency in the deployment of the interns, we identified the period of their absence (1^st^ April 2023 to 6^th^ August 2023) and presence (the rest of the period).

**Covariates:** We extracted data on the woman’s age in years, parity, area of residence, gestational age assessed using the woman’s last normal menstrual period, woman’s referral status, the indication of emergency CS, calendar day of operation, work shift of the healthcare professionals, and type of anaesthesia.

### Sample size

Using a two-proportion formula, we calculated a minimum required sample size of 760 participants. Assuming that 31% of emergency CS performed by senior doctors were prolonged DDI (>75 minutes) according to a previous study in Uganda(4), this sample size was adequate to detect a 32% relative difference in the proportion of prolonged DDI for emergency CS done by juniors compared to senior doctors, with a two-sided significance level of 0.05 and 80% power.

### Statistical methods

We conducted statistical analyses using STATA 18 (Texas, USA). Descriptive statistics included means and standard deviations for normally distributed continuous variables, medians and interquartile ranges for skewed distributions, and frequencies and percentages for categorical variables.

For univariable analysis, we employed chi-squared tests and logistic regression to assess associations between exposures, covariates, and prolonged DDI, presenting percentages, crude odds ratios (OR), 95% confidence intervals (CI), and likelihood ratio test (LRT) p-values.

We performed minimal model analysis using logistic regression, considering woman’s age as an a priori confounder. We fitted three initial models, each including one main exposure, age, the DDI outcome, and potential confounders individually. We identified important confounders by examining changes in exposure odds ratios.

For multivariable analysis, we used causal modelling and a forward modelling approach. We constructed three final models, each including one main exposure, age, and important confounders identified from the minimal-model analysis. We assessed multicollinearity by comparing natural log standard errors between initial and full models.

We explored effect modification by emergency CS indication, comparing final adjusted models with and without interaction terms using LRT. We reported stratum-specific odds ratios for each indication category. For missing data, we assessed data completeness and patterns of missingness. Our primary analysis used complete case analysis and did not adjust for variables with substantial missing data (>5%) as potential confounders to minimise bias. In sensitivity analyses, the impact of additional adjustment for the variables with >5% missing data was examined. All statistical tests were two-sided, with significance at p<0.05.

### Ethical approval

Ethical approval was obtained from the St. Mary’s Hospital Lacor Research Ethics Committee, reference number LACOR-2023-272. Permission to conduct the study at St. Mary’s Hospital Lacor was granted by the hospital administration. Informed consent was waived off by the ethics committee given the retrospective nature of the study and confidentiality of patient information was strictly maintained throughout the research process.

## Results

### Participants

Of 772 women who underwent emergency CS between 6^th^ September 2022 and 1^st^ June 2024, 760 (98.4%) met the inclusion criteria for our study. We excluded 0.4% (3/772) of the women delivering at gestational age <24 weeks and 1.2% (9/772) women with multiple pregnancy. The final cohort of 760 women were followed up from enrolment when an emergency CS was prescribed to delivery of the baby. There was no loss to follow-up (Fig 1).

**Fig 1.**
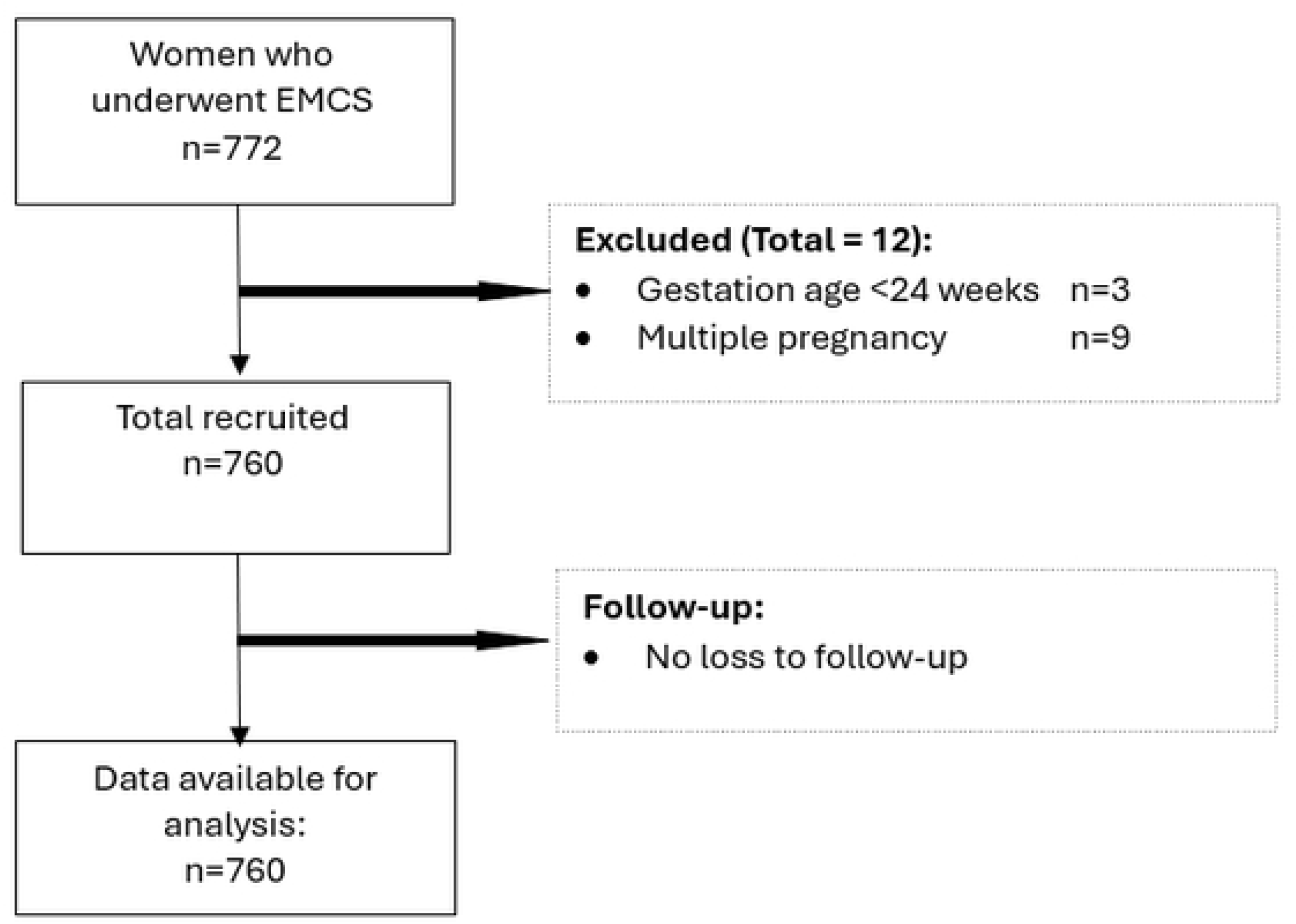
Flow diagram of participant selection.

### Participant characteristics

The mean age of the women was 25 years (standard deviation, SD: 6.0), with the majority, 56.7% (n=431), of the women in the 20-29 years age group. The median parity of the women was 1 (interquartile range, IQR: 1-3) and 35.3% (268) were multiparous. The median gestation age was 39 weeks (IQR: 38-40) with most women, 66.5% (n=410), delivering at term. A majority of women: 59.6% (n=453) were referred from other health facilities to St. Mary’s Hospital Lacor; 70.1% (n=533) resided in non-city districts of Uganda; and 83.3% (n=633) underwent emergency CS under spinal anaesthesia. Twenty-four babies (3.2%) were stillbirths while 18.7% (n=142) of the babies born had birth asphyxia. Nearly four in ten (n=288) had emergency CS due to previous CS in active labour (Table 1).

**Table 1:** Characteristics of women and their newborn outcome who underwent emergency CS at St. Mary’s Hospital Lacor between 6^th^ September 2023 and 1^st^ June 2024 (N=760).

| Characteristics | n | (%)* |
| --- | --- | --- |
| Surgeon's cadre□ |  |  |
| Junior | 169 | 25.2 |
| Medical officer | 471 | 70.1 |
| Specialist | 32 | 4.8 |
| Theatre |  |  |
| Main | 582 | 76.6 |
| Labour-suite | 178 | 23.4 |
| Presence of interns□ |  |  |
| Present | 569 | 74.9 |
| Absent | 189 | 24.9 |
| Point of pre-operative delay |  |  |
| None | 138 | 18.2 |
| Ward | 186 | 24.5 |
| Theatre | 186 | 24.5 |
| Ward+theatre | 250 | 32.9 |
| DDI (minutes) |  |  |
| Median(IQR) | 51 | 36-67 |
| <60 | 486 | 64.0 |
| ≥60 | 274 | 36.0 |
| Woman's age (years) |  |  |
| Mean(SD) | 25.0 | 6.0 |
| <20 | 153 | 20.1 |
| 20-29 | 431 | 56.7 |
| ≥30 | 176 | 23.2 |
| Gestational age (weeks)□ |  |  |
| Median(IQR) | 39 | 38-40 |
| Preterm | 95 | 12.5 |
| Term | 410 | 66.5 |
| Post-term | 57 | 7.5 |
| Parity□<sup>a</sup> |  |  |
| Median(IQR) | 1 | 1-3 |
| Nulliparous | 176 | 23.2 |
| Primiparous | 213 | 28.0 |
| Multiparous | 268 | 35.3 |
| Grand multiparous | 100 | 13.2 |
| Referral status |  |  |
| No | 307 | 40.4 |
| Yes | 453 | 59.6 |
| Work shift** |  |  |
| Morning | 237 | 31.2 |
| Afternoon | 214 | 28.2 |
| Night | 309 | 40.7 |
| Calendar day□ |  |  |
| Weekday | 488 | 64.2 |
| Weekend/public holiday | 270 | 35.5 |
| Residence |  |  |
| City | 227 | 29.9 |
| Non-city | 533 | 70.1 |
| Indication of emergency CS |  |  |
| Previous CS | 288 | 37.9 |
| Foetal distress | 125 | 16.5 |
| Obstructed labour | 148 | 19.5 |
| Others | 199 | 26.2 |
| Type of anaesthesia□ |  |  |
| Spinal anaesthesia | 633 | 83.3 |
| General anaesthesia | 38 | 5.0 |
| Newborn outcome |  |  |
| Stillbirth | 24 | 3.2 |
| Birth asphyxia | 142 | 18.7 |
| Normal | 594 | 78.2 |

**Abbreviations:** DDI, decision to delivery interval; CS, caesarean section; SD, standard deviation; IQR, inter-quartile range.
<sup>a</sup>Parity refers to previous pregnancies delivered alive or dead after 24 weeks of gestation grouped into nulliparous, primiparous, multiparous, and grand multiparous (0, 1, 2-4 and ≥5 respectively)
□ Missing data: Gestational age 198 (26.1%), parity 3 (0.4%), type of anaesthesia 89 (11.7), surgeon cadre 88 (11.6%), calendar day 2 (0.3%) and presence of interns 2 (0.3).
\*Percentages may not add up to 100.0% due to rounding and missing data.
\*\*Work shift – Morning (08:00-13:59 hours), afternoon (14:00-19:59 hours), and night (20:00-07:59 hours).

#### Exposures and outcome

A majority, 74.9% (n=503) of the women were operated on by senior doctors and 76.6% (n=582) were operated on in the hospital’s main theatre. The median DDI was 51 minutes (IQR: 36-67) with nearly two-thirds, 64.0% (n=486) of women having delivery in less than 60 minutes. Three quarters of emergency CS deliveries 74.9% (n=569) occurred during a period when intern healthcare professionals were available (Table 1).

### Crude analysis

#### Exposures-DDI association

Emergency CS performed by junior doctors had significantly higher odds of prolonged DDI compared to those performed by senior doctors, with 51.5% (87/169) of the junior-led emergency CS experiencing delays versus 31.0% (156/503) for senior-led emergency CS (OR: 2.36, 95% CI: 1.65-3.37; p<0.0001). The location of operating theatre showed no significant association with prolonged DDI, as similar proportions of women experienced delays in the main theatre (36.1%; 210/582) and labour-suite (36.0%; 64/178) (OR: 0.99, 95% CI: 0.70-1.41;p=0.98). Regarding the presence of intern healthcare professionals, prolonged DDI was slightly more frequent when interns were present (38.0%; 216/569) than absent (30.2%; 57/189), though the association was weak (OR:0.71, 95% CI: 0.50-1.01; p=0.05) (Table 2).

**Table 2:** Surgeon’s cadre, use of labour-suite theatre, presence of intern healthcare professionals, and other covariates and their unadjusted association with prolonged DDI estimated by logistic regression (N=760).

| Variable |  | N (%)<sup>*</sup> | Prolonged DDI<br>n (%)<sup>*</sup> | OR<br>(95% CI) | p-value |
| --- | --- | --- | --- | --- | --- |
| Surgeon's cadre□ | Senior doctor | 503 (74.9) | 156 (31.0) | 1 | <0.0001 |
|  | Junior | 169 (25.2) | 87 (51.5) | 2.36 (1.65-3.37) |  |
| Theatre | Main | 582 (76.6) | 210 (36.1) | 1 | 0.98 |
|  | Labour-suite | 178 (23.4) | 64 (36.0) | 0.99 (0.70-1.41) |  |
| Presence of interns□ | Present | 569 (74.9) | 216 (38.0) | 1 | 0.05 |
|  | Absent | 189 (24.9) | 57 (30.2) | 0.71 (0.50-1.01) |  |
| Point of pre-operative delay<sup>a</sup> | Theatre-related | 186 (24.5) | 62 (33.3) | 1 | <0.0001 |
|  | Ward-related | 186 (24.5) | 69 (37.1) | 1.18 (0.77-1.81) |  |
|  | Ward+theatre | 250 (32.9) | 143 (57.2) | 2.67 (1.80-3.97) |  |
| Woman's age (years) | <20 | 153 (20.1) | 57 (37.3) | 1.01 (0.69-1.47) | 0.51 |
|  | 20-29 | 431 (56.7) | 160 (37.1) | 1 |  |
|  | ≥30 | 176 (23.2) | 57 (32.4) | 0.81 (0.56-1.18) |  |
| Gestational age (weeks)□ | Preterm | 95 (12.5) | 42 (44.2) | 1.37 (0.87-2.16) | 0.39 |
|  | Term | 410 (66.5) | 150 (36.6) | 1 |  |
|  | Post-term | 57 (7.5) | 21 (36.8) | 1.01 (0.57-1.80) |  |
| Parity□<sup>b</sup> | Nulliparous | 176 (23.2) | 80 (45.5) | 1 | 0.005 |
|  | Primiparous | 213 (28.0) | 66 (31.0) | 0.54 (0.36-0.82) |  |
|  | Multiparous | 268 (35.3) | 100 (37.3) | 0.71 (0.49-1.05) |  |
|  | Grand multiparous | 100 (13.2) | 27 (27.0) | 0.44 (0.26-0.76) |  |
| Referral status | No | 307 (40.4) | 111 (36.2) | 1 | 0.96 |
|  | Yes | 453 (59.5) | 163 (36.0) | 0.99 (0.73-1.34) |  |
| Work shift<sup>**</sup> | Morning | 237 (31.2) | 70 (29.5) | 1 | 0.02 |
|  | Afternoon | 214 (28.2) | 78 (36.5) | 1.37 (0.92-2.03) |  |
|  | Night | 309 (40.7) | 126 (40.8) | 1.64 (1.15-2.35) |  |
| Calendar day□ | Weekday | 488 (64.2) | 170 (34.8) | 1 | 0.36 |
|  | Weekend/public holiday | 270 (35.5) | 103 (38.2) | 1.15(0.85-1.57) |  |
| Residence | City | 227 (29.9) | 88 (38.8) | 1 | 0.31 |
|  | Non-city | 533 (70.1) | 186 (34.9) | 0.85 (0.61-1.17) |  |
| Indication of emergency CS | Previous CS | 288 (37.9) | 105 (36.5) | 1 | 0.48 |
|  | Foetal distress | 125 (16.5) | 52 (41.6) | 1.24 (0.81-1.91) |  |
|  | Obstructed labour | 148 (19.5) | 50 (33.8) | 0.89 (0.59-1.35) |  |
|  | Others | 199 (26.2) | 67 (33.67) | 0.88 (0.61-1.29) |  |
| Type of anaesthesia□ | Spinal | 633 (83.3) | 235 (37.1) | 1 | 0.04 |
|  | General | 38 (5.0) | 8 (21.1)) | 0.45 (0.20-1.00) |  |

**Abbreviations:** DDI, decision to delivery interval; CS, caesarean section; OR, odds ratio; CI, confidence interval.
<sup>a</sup>Women without any delays documented 138 (18.2%) excluded to overcome zero cells when assessing point of delay-prolonged DDI association
<sup>b</sup>Parity refers to previous pregnancies delivered alive or dead after 24 weeks of gestation grouped into nulliparous, primiparous, multiparous, and grand multiparous (0, 1, 2-4 and ≥5 respectively)
□Missing data: Gestational age 198(26.1%), parity 3(0.4%), type of anaesthesia 89(11.7), surgeon cadre 88(11.6%).
\*Percentages may not add up to 100.0% due to rounding and missing data.
\*\*Work shift – Morning (08:00-13:59 hours), afternoon (14:00-19:59 hours), and night (20:00-07:59 hours).

#### Covariate-DDI association

Prolonged DDI was more frequent among women who experienced combined ward and theatre-related delays compared to those with delays confined to a single area. Nulliparous women had a higher proportion of prolonged DDI compared to primiparous, multiparous, and grand multiparous women. The timing of surgery also influenced delays, with higher proportions of prolonged DDI observed during afternoon and night shifts than during morning shifts. Additionally, women operated under spinal anaesthesia were more likely to experience prolonged DDI than those who underwent general anaesthesia (Table 2).

### Confounder selection

We identified key confounders for multivariable analysis: for the surgeon’s cadre-DDI relationship, these included work shift, calendar day, presence of juniors, gestational age, and parity; for the theatre-DDI relationship, they included work shift, calendar day, presence of juniors, area of residence, gestational age, and parity. Apart from maternal age (a forced variable), no other confounders were identified for the presence of interns based on expert knowledge. Point of pre-operative delay and type of anaesthesia were considered mediators in the surgeon’s cadre-DDI pathway and were not adjusted for.

### Adjusted analysis

After adjusting for confounders, emergency CS performed by junior doctors remained significantly associated with prolonged DDI compared to senior doctors, with adjusted odds nearly double (OR: 2.07, 95% CI: 1.38–3.10; p=0.0004). The choice of operating theatre showed no significant association with prolonged DDI after adjustment, with women operated in the labour-suite theatre having similar odds of delays as those in the main theatre (OR: 0.89, 95% CI: 0.61–1.28; p=0.52). For the presence of intern healthcare professionals, the adjusted analysis showed weak evidence of an association, with slightly lower odds of prolonged DDI during periods of intern absence (OR: 0.71, 95% CI: 0.50–1.02; p=0.06) (Table 3).

**Table 3:** Adjusted estimates of the odds ratio for association between surgeon’s cadre, operating theatre choice, and presence of intern healthcare professionals, and prolonged DDI estimated by logistic regression

| Variable |  | OR<br>(95% CI) | p-value | OR<br>(95% CI) | p-value |
| --- | --- | --- | --- | --- | --- |
| Surgeon's cadre | Senior doctor | 1 | <0.0001 | 1 | 0.0004 |
|  | Junior | 2.35 (1.64-3.36)<sup>a</sup> |  | 2.07 (1.38-3.10)<sup>c</sup> |  |
| Theatre | Main | 1 | 0.91 | 1 | 0.52 |
|  | Labour-suite | 0.98 (0.69-1.39)<sup>a</sup> |  | 0.89 (0.61-1.28)<sup>d</sup> |  |
| Presence of interns | Present | 1 | 0.05 | 1 | 0.06 |
|  | Absent | 0.71 (0.50-1.01)<sup>b</sup> |  | 0.71 (0.50-1.02)<sup>e</sup> |  |

**Abbreviations:** DDI, decision-to-delivery interval; OR, odds ratio; CI, confidence interval.
□ Likelihood ratio p-values
<sup>a</sup>OR adjusted for age
<sup>b</sup>Crude OR
<sup>c</sup>OR adjusted for age, parity group, presence of juniors, calendar day of the week, and work shift (N=667).
<sup>d</sup>OR adjusted for age, parity group, indication of emergency caesarean section, area of residence, and calendar day of the week (N=755).
<sup>e</sup>OR adjusted for age (N=670).

### Effect modification by indication of emergency CS

There was no evidence that the association between surgeon’s cadre, choice of operating theatre, and the presence of intern healthcare professionals and the odds of prolonged DDI varied by the indications of the emergency CS (p=0.83, p=0.47, and p=0.88 respectively) (S1 Table in the supporting information).

### Sensitivity analysis

On sensitivity analysis: inclusion of gestational age in the final model for the adjusted association between surgeon’s cadre and choice of operating theatre and the odds of prolonged DDI association yielded similar results (OR: 2.00, 95% CI: 1.30-3.09, p=0.002, N=487, and OR: 0.91, 95% CI: 0.59-1.41, p=0.67, N=559 respectively) (S2 Table in the supporting information).

## Discussion

Our study provides strong evidence that emergency CS performed by junior doctors were associated with significantly higher odds of a prolonged DDI compared to those performed by senior doctors. We also found no significant difference in the odds of prolonged DDI between emergency CS performed in the distant main hospital theatre versus the labour-suite theatre. Furthermore, the presence of intern healthcare professionals showed weak evidence of an association with prolonged DDI, with slightly lower odds of prolonged DDI during periods when interns were absent. We did not find any evidence that the associations between surgeon’s cadre, operating theatre location, and intern presence and prolonged DDI varied significantly by the indication for emergency CS.

The association between junior doctors performing emergency CS and prolonged DDI highlights the important role that surgeon experience plays in timely emergency obstetric care. This finding aligns with previous research suggesting that less experienced surgeons are more likely to encounter delays in performing emergency procedures (20,23). This finding highlights the need for robust training, mentorship, and supervision for junior doctors to enhance their efficiency and decision-making in emergency CS situations (24). The absence of a significant impact of operating theatre location on DDI is a novel finding. Despite the distance between the labour-suite and the main theatre, our result suggests that effective coordination and organisation between the two theatres at St. Mary’s Hospital Lacor may mitigate the impact of this distance on DDI. Factors such as the readiness of the theatre staff, the timely availability of anaesthesia, and overall operational efficiency may play a more critical role in determining DDI than the proximity of the operating theatre. The weak association between the presence of intern healthcare professionals and DDI suggests that, while interns contribute to the workforce, their presence does not have a substantial impact on DDI. This could indicate that other factors, such as the specific tasks interns are involved in, the level of supervision they receive, and the overall team coordination, might play a more significant role in determining the timeliness of emergency CS.

Our study has several strengths. First, the large sample size and high data completeness for most variables strengthen the reliability of our findings. Moreover, the cohort design allowed for the examination of multiple exposures, adjusting for a range of potential confounders, and provided a comprehensive assessment of the factors influencing DDI in our setting. Our study also adds an unexplored aspect of studying the impact of proximity of the operating theatre to labour-suite on the DDI, an intervention not yet investigated in previous studies. However, several limitations should be considered in the interpretation of these results. Despite our efforts to adjust for known confounders, unmeasured factors, such as socio-economic status or institutional constraints, may have biased our results. Additionally, as a single-centre study conducted at a private tertiary hospital, the findings may not be fully generalisable to other healthcare settings, particularly primary healthcare and public health facilities. Lastly, while DDI is an important process indicator, our study did not directly assess maternal or neonatal outcomes.

Our findings have several important implications for improving emergency obstetric care in resource-limited settings. First, there is critical need for enhanced training and supervision for junior doctors performing emergency CS through programs such as structured mentorship or simulation-based training that could improve their efficiency and reduce DDI, ultimately leading to better outcomes for mothers and babies (25,26). Second, healthcare facilities should continue to focus on improving theatre readiness, team coordination, and timely communication to minimise delays, regardless of the physical distance between theatre locations. Future research should focus on the direct impact of prolonged DDI on maternal and neonatal outcomes, as well as evaluate interventions like mentorship or simulation-based training to reduce delays. Additionally, understanding the barriers junior doctors face in delivering timely emergency CS will provide valuable insights for improving emergency obstetric care in similar settings.

## Conclusions

This study provides evidence of the associations between surgeon’s cadre, theatre location, and the presence of intern healthcare professionals with prolonged DDI in emergency CS at a tertiary hospital in Northern Uganda. Our findings highlight the importance of addressing surgeon experience and system-level delays in emergency obstetric care. By focusing on enhancing training, improving theatre processes, and optimising team dynamics, healthcare facilities in resource-limited settings can improve the timeliness of emergency CS, potentially leading to better maternal and neonatal outcomes(27).

## Data Availability

The data underlying the findings of this study are available upon request. Interested parties may contact the corresponding author to access the data.

## Acknowledgements

We would like to extend our heartfelt gratitude to the participants. Our sincere thanks go to the doctors, nurses, midwives, anaesthesia staff, and the auxiliary staff at St. Mary’s Hospital Lacor for their invaluable support and collaboration throughout the data collection process. We are also deeply appreciative of the hospital administration for permitting us to conduct this research and for their continuous assistance. This study would not have been possible without the collective efforts of everyone involved.

## Supporting information

### Supporting information caption

**S1 Table.** Content and legend are strongly recommended.

**S2 Table.** Content and legend are optional.

